# Virtual transitional pain service delivered via telehealth is effective in preventing new and persistent opioid use amongst post-surgical spine patients

**DOI:** 10.1101/2023.08.18.23294272

**Authors:** Maryam Hussain, Beau Norgeot, Ahmed Zaafran, Jessica Stark, John Caridi, Albert Fenoy, Evan Pivalizza

## Abstract

Opioid dependence is a national crisis, with 30 million patients annually at risk of becoming persistent opioid users after receiving opioids for post-surgical pain management. Translational Pain Services (TPS) demonstrate effectiveness for behavioral health improvements but its effectiveness in preventing persistent opioid use is less established, especially amongst opioid exposed patients. Prohibitive costs and accessibility challenges have hindered TPS program adoption. To address these limitations, we designed and implemented a remote telehealth TPS protocol focusing on preventing continued opioid use while improving behavioral health. Licensed therapists trained in the opioid-tapering CBT protocol delivered sessions reimbursed through standard payer reimbursement. Our prospective study evaluated the protocol’s effectiveness on preventing persistent opioid use and behavioral health outcomes amongst both opioid naïve and exposed patients. In an opioid-naive patient cohort (n=67), 100% completely tapered off opioids, while in an opioid-exposed cohort (n =19) 52% completely tapered off opioids, demonstrating promising results. In both cohorts, we observed significant improvements in behavioral health scores, including pain. This opioid-tapering digital TPS is effective, adoptable, and incurs no out-of-pocket cost for healthcare systems. We provide the opioid-tapering CBT protocol in the supplement to facilitate adoption. *Trial Registration* Impact of Daily, Digital and Behavioral Tele-health Tapering Program for Perioperative Surgical Patients Exposed to Opioids and Benzodiazepines registered at clinicaltrials.gov, NCT04787692. https://clinicaltrials.gov/ct2/show/NCT04787692?term=NCT04787692&draw=2&rank=1

## Introduction

The opioid epidemic has claimed the lives of over 500,000 people in America.^1^ Far from affecting only a marginalized community, many patients are initially exposed to opioids after a surgical or traumatic event. This epidemic has been further accelerated by the COVID pandemic, with a 46% increase in opioid-related overdoses and 13% increase in overdose-related deaths from 2019 to 2020.^2^ There are 80 million major surgeries per year in the United States and between 49-95% of those post-surgical patients are discharged with an opioid prescription.^3–6^ This puts approximately 30 million new Americans at an increased risk of persistent opioid use and the severe adverse events associated with it every year.^7^ However, while there are no direct estimates to indicate how many patients receive an opioid tapering care plan, extant literature suggests that most patients are discharged with the expectation they will taper on their own.^8,9^

Post-surgical pain is a major contributor to continued opioid use.^10^ Spine surgery is associated with new and persistent opioid use, with 13% of opioid naive patients continuing to use opioids for more than 90-days after surgery.^11,12^ Risk of continued opioid use varies based on opioid exposure prior to surgery. As such, opioid exposed patients, or those who have been using opioids to manage pain prior to surgery, may have different postoperative pain expectations and management challenges compared to opioid naive patients, or those who have *not* been using opioids for pain management prior to surgery.^13^ Furthermore, opioid use is associated with poorer behavioral health outcomes (e.g., depression, anxiety, and quality of life) and increased healthcare costs.^14^

In particular, opioid prescription use, misuse and side effects comprise a significant portion of postoperative health care costs, with some estimates showing that a person with continued opioid use has annual healthcare costs $14,054–$20,546 greater than a person who does not use opioids.^15^ Findings from an IBM^R^ Marketscan^R^ insurance claims database indicate that amongst orthopedic and spine surgery patients, almost 10% have an opioid-related hospitalization and 18% have an emergency room (ER) visit within 90 days after surgery, which can account for around $32,000 per event in healthcare costs.^16^ Similar findings from a separate study using IBM^R^ MarketScan^R^ research databases support these findings.^17^

Most surgical patients do not wish to be on opioid therapy,^18^ are fearful of dependency, and want to be involved in their tapering process.^19^ Transitional pain services (TPS) have demonstrated success in helping patients manage pain after surgery, as well as improve behavioral health outcomes.^20,21^ However, TPS’s efficacy in preventing persistent opioid use seems less established. In one study, 27% (n =19) of the patients were still using opioids three months postoperatively, with over half of this cohort (ten of the 19 patients) being opioid naïve preoperatively.^20^ In another study, opioid tapering results were even more varied: 46% (n = 51) of opioid naïve patients had completely tapered, 45% (n =5 0) had reduced opioid usage, and 8% (n = 9) actually had increased their dosage within the six months after surgery. Tapering results at the six-month postoperative mark were less successful amongst opioid exposed patients: 25% (n = 35) tapered completely, 56% (n = 77) had reduced opioid usage, and 19% (n =27) had increased their dosage.^21^ Beyond the limitations in completely tapering patients off opioids, brick-and-mortar TPS has not been widely adopted,^21^ potentially due to cost and accessibility challenges.^22,23^

Thus, to address these limitations of brick-and-mortar TPS, we designed a remote telehealth TPS protocol focusing on preventing continued opioid use while improving behavioral health. We incorporated an opioid-tapering CBT protocol as CBT has also been demonstrated to improve post-surgical pain management, behavioral health, and emotional wellbeing.^25^ Licensed therapists trained in this new protocol digitally delivered sessions to patients, which were paid through standard payer reimbursements, effectively removing cost and accessibility hurdles. We then assessed the feasibility and effectiveness of this new TPS protocol through a dual-arm prospective study of opioid naïve and exposed patients undergoing elective spinal surgery at an academic tertiary care hospital. The primary outcomes assessed were participant engagement, opioid tapering, and self-reported pain, quality of life, anxiety, and depression. We did not directly assess cost-savings, but we did track postoperative emergency room visits and hospitalizations, which are the major drivers of healthcare cost associated with the opioid epidemic.^17^

## Results

After screening 298, 150 participants were enrolled. Baseline information on participants is presented in Table 1. The majority identified as female (n = 89, 59·3%). Race distribution was diverse with 59·2% identifying as White (n = 89), 20·0% African American/Black (n = 30), 19·3% as Hispanic/Latino (n = 29), and 1·3% as Other (n = 2). One hundred and one of participants were opioid naive, and 49 were opioid exposed. Opioid exposed participants had worse baseline behavioral health scores than the opioid naive group on all baseline behavioral health scores (all p < 0·001) (Table 1 and Supplementary Table 1).

**Table 1.** Patient Preoperative Characteristics.

| Characteristic | n | Mean (SD) or % |
| --- | --- | --- |
| Age (years) |  | 56.6 (12.8) |
| Sex |  |  |
| Male | 62 | 41.3% |
| Female | 88 | 58.7% |
| Other | 0 | 0.0% |
| Ethnicity |  |  |
| Black/African American | 30 | 20.0% |
| Hispanic/Latino | 29 | 19.3% |
| White | 89 | 59.3% |
| Other | 2 | 1.3% |
| Preoperative opioid use |  |  |
| Yes (Opioid Exposed) | 49 | 32.70% |
| No (Opioid Naive) | 101 | 67.30% |
| Preoperative Behavioral Health |  |  |
| Pain<sup>a</sup> |  | 6.6 (2.5)*<sup>a</sup> |
| QoL<sup>b</sup> |  | 57.5 (27.7)*<sup>b</sup> |
| Functioning<sup>c</sup> |  | 73.8 (11.6)*<sup>c</sup> |
| Depression<sup>d</sup> |  | 7.7 (6.4)*<sup>d</sup> |
| Anxiety<sup>e</sup> |  | 7.3 (5.9)*<sup>e</sup> |
| Symptoms<sup>f</sup> |  | 29.5 (19.8)*<sup>f</sup> |

<sup>a</sup>Visual Analog Scale (numerical rating 0-10)
<sup>b</sup>European Quality of Life EQ5D5L composite score (0-100)
<sup>c</sup>Global Assessment of Functioning (numerical rating 0-100)
<sup>d</sup>Patient Health Questionnaire-9 total score
<sup>e</sup>Generalized Anxiety Disorder-7 total score
<sup>f</sup>Edmonton Symptoms Assessment Survey total score
\*<sup>a</sup> statistically significantly higher pain symptoms for opioid exposed, $F(1,149) = 19.9$ , $p < .001$ , Cohen's $f = .35$
\*<sup>b</sup> statistically significantly lower QoL for opioid exposed, $F(1,149) = 22.4$ , $p < .001$ , Cohen's $f = .38$
\*<sup>c</sup> statistically significantly lower functioning for opioid exposed, $F(1,149) = 25.5$ , $p < .001$ , Cohen's $f = .40$
\*<sup>d</sup> statistically significantly higher depression symptoms for opioid exposed, $F(1,149) = 20.9$ , $p < .001$ , Cohen's $f = .36$
\*<sup>e</sup> statistically significantly higher anxiety symptoms for opioid exposed, $F(1,149) = 11.1$ , $p = .001$ , Cohen's $f = .26$
\*<sup>f</sup> statistically significantly higher overall symptoms for opioid exposed, $F(1,149) = 20.9$ , $p < .001$ , Cohen's $f = .36$

Of the 150 consented cohort, 66·0% of the naive and 55·0% of the exposed participants engaged with aggregate engagement of 63·0% (Figure 1). Engagement was not statistically associated with age, sex, ethnicity, or preoperative behavioral health scores (Supplementary Table 2).

**Figure 1.**
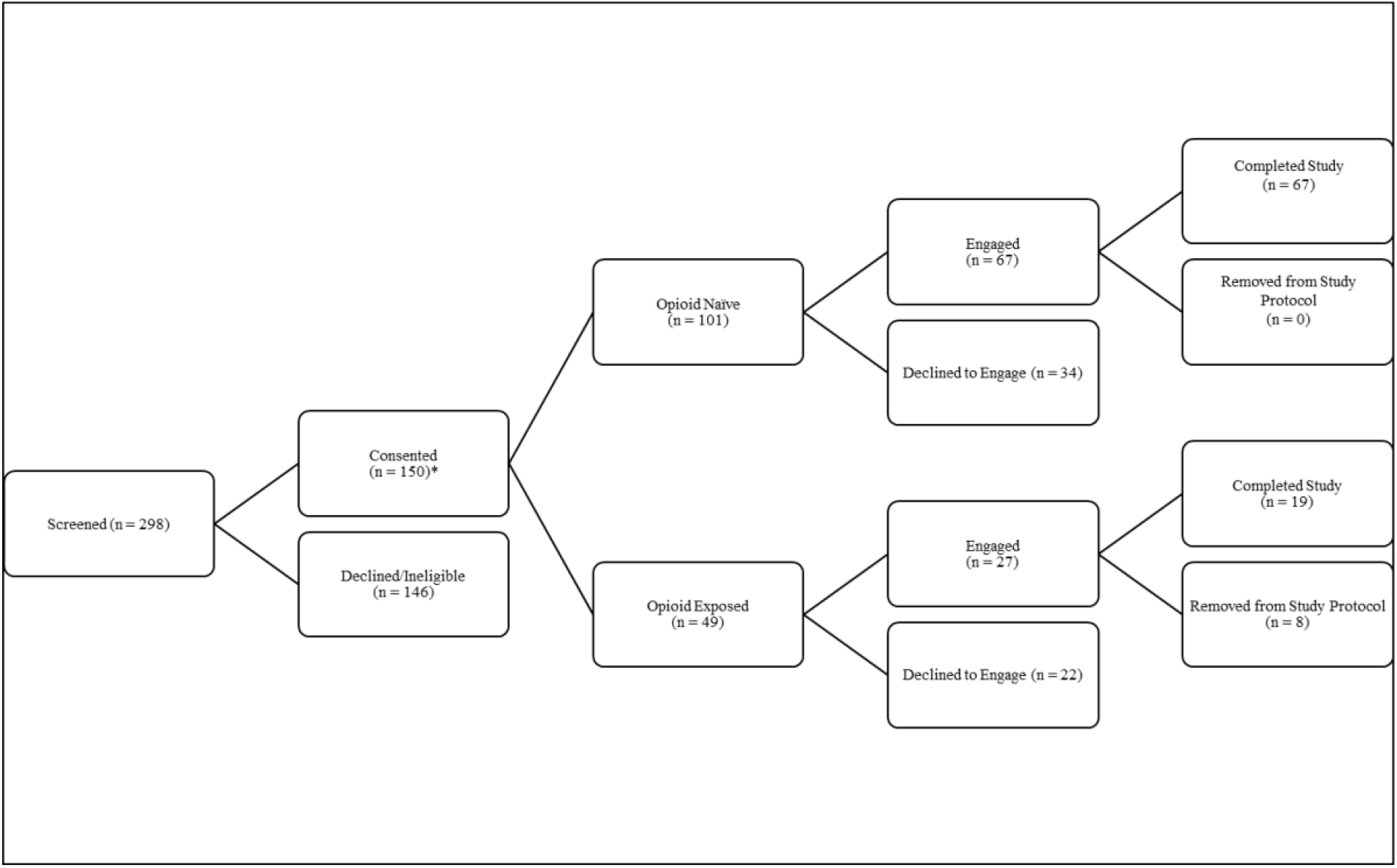
CONSORT Diagram. * n = 152 consented; however, n = 2 were promptly removed from consent procedures due to cancellation of surgical procedure

There were significant differences in postoperative opioid dosages at the initial therapy session between the opioid naive versus exposed groups (26 MME vs 42 MME), *F*(1, 85) = 4·7, p = ·03, Cohen’s *f* = ·21. Both groups saw clinically and statistically significant reductions in dosage at their final postoperative appointment compared to postoperative baseline. The exposed group reduced MME consumption by 65·0% (avg 14 MME), with 53·0% of participants tapering completely off their postoperative opioids (0 MME). Finally, 100% of the opioid naive arm patients tapered completely off their postoperative opioids (0 MME). Opioid naïve patients statistically significantly tapered 100% of postoperative opioids compared to opioid exposed patients, *F*(1, 85) = 35·0, p < ·001, Cohen’s *f* = .63.

By the final postoperative appointment, both groups had statistically and clinically significant improvements in all behavioral health metrics from initial postoperative scores, which are presented as median with interquartile range (IQR). The opioid naive arm (Figure 2) had 43·7% improvement in pain (baseline VAS = 5·5, IQR = 4·3; final VAS = 3·0, IQR = 2·0), 17·5% improvement in QoL (baseline EQ5D5L= 67·8, IQR = 27·5; final EQ5D5L= 77·5, IQR = 13·6), 6·3% improvement in functioning (baseline GAF = 75·0, IQR = 15·0; final GAF = 85·0, IQR = 9·0), 27·0% improvement in common symptoms (baseline ESAS = 26·0, IQR = 16·0; final ESAS = 20·0, IQR = 23·0), 66·7% improvement in anxiety (baseline GAD7 = 3·0, IQR = 6·0; final GAD7 = 0·0, IQR = 3·0), and 72·7% improvement in depression (baseline PHQ9 = 4·0, IQR = 8·0; final PHQ9 =0·0, IQR = 4·0). See Supplementary Table 3 for Wilcoxon signed rank test statistics.

**Figure 2.**
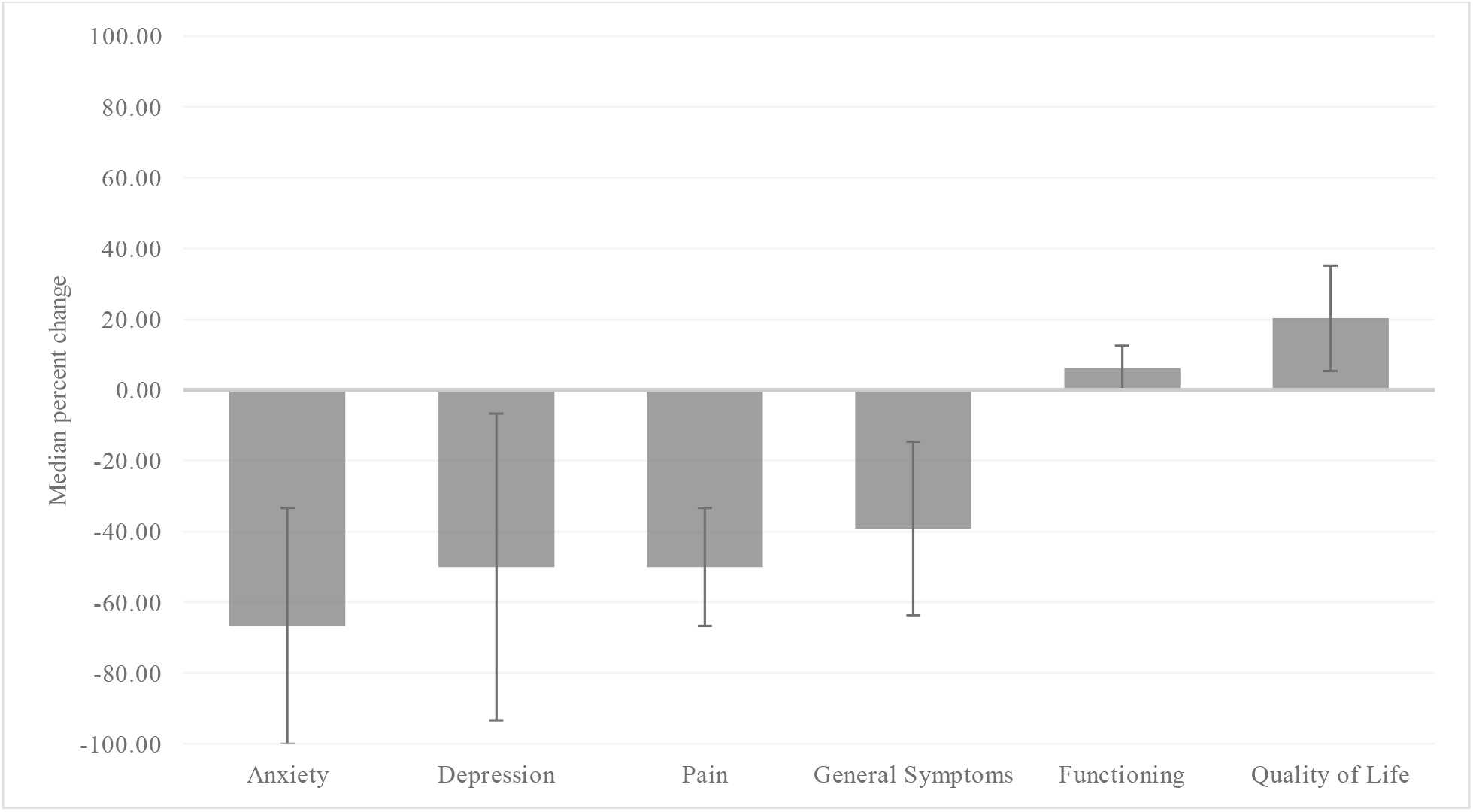
Behavioral health changes within 90-days for the opioid naive arm. *Note*.Reduced scores in anxiety, depression, pain, and general symptoms indicate improvement. Increased scores in functioning and quality of life indicate improvement. All behavioral health median changes were statistically significantly different from the first appointment to the final appointment (*p* <·001).

The opioid exposed arm (Figure 3) had 33·3% improvement in pain (baseline VAS = 6·0, IQR = 5·0; final VAS = 4·0, IQR = 2·0), 100·0% improvement in QoL (baseline EQ5D5L= 44·0, IQR = 58·3; final EQ5D5L= 79·6, IQR = 19·4), 9·4% improvement in functioning (baseline GAF = 72·0, IQR = 12·0; final GAF = 80·0, IQR = 15·0), 34·6% improvement in common symptoms (baseline ESAS = 33·0, IQR = 5·0; final ESAS = 21·0, IQR = 25·5), 82·1% improvement in anxiety (baseline GAD7 = 9·5, IQR = 12·0; final GAD7 = 2·0, IQR = 4·0), and 57·3% improvement in depression (baseline PHQ9 = 11·5, IQR = 11·5; final PHQ9 = 3·0, IQR = 5·5). See Supplementary Table 4 for Wilcoxon signed rank test statistics.

**Figure 3.**
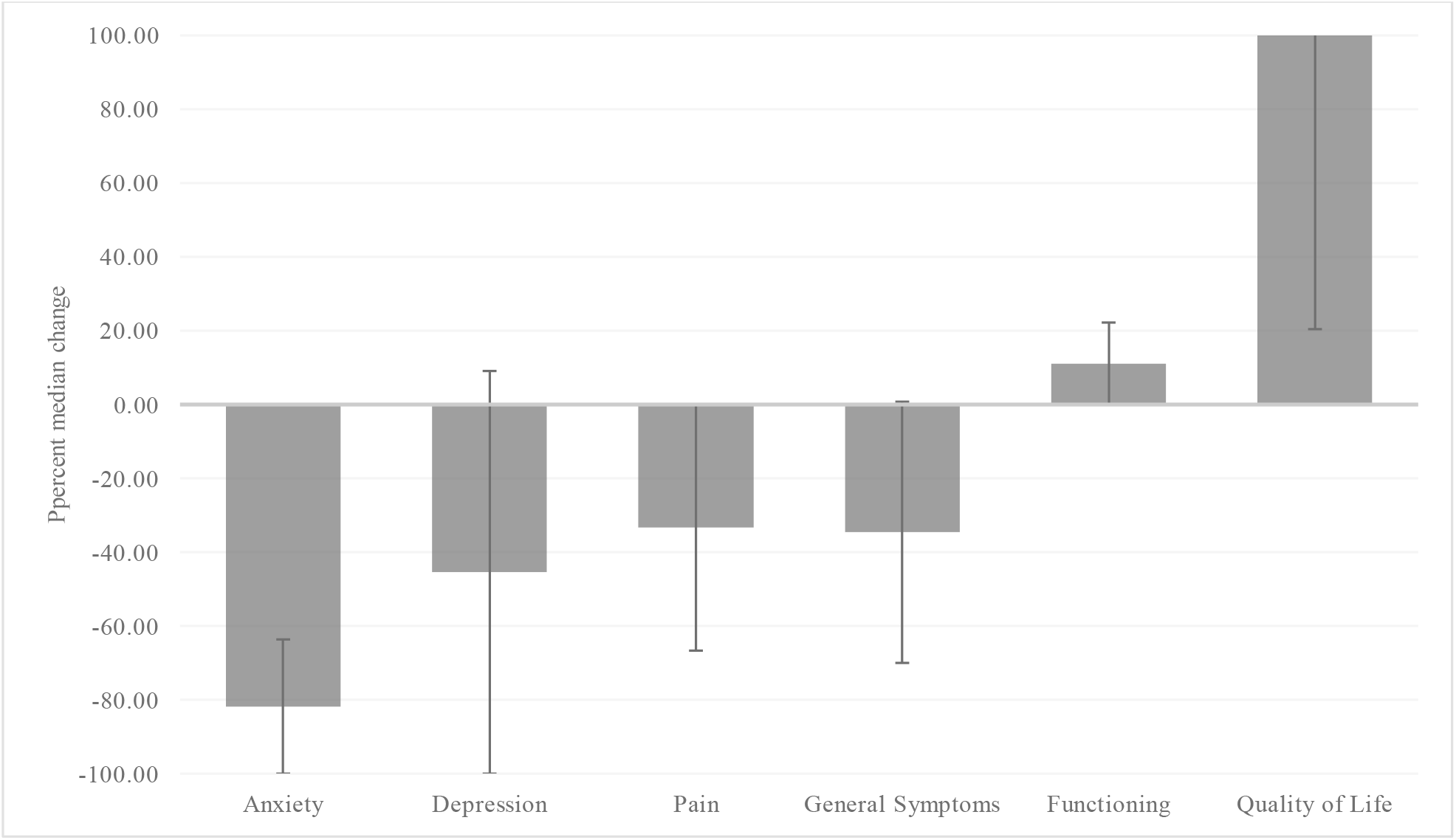
Behavioral health changes with statistical significance for the Opioid Exposed Arm. *Note*. Reduced scores in anxiety, depression, pain, and general symptoms indicate improvement. Increased scores in functioning and quality of life indicate improvement. All behavioral health changes were statistically significantly different from the first appointment to the final appointment (*p* <·05).

There were no high-cost healthcare utilization events, particularly postoperative ER visits and/or re-hospitalizations, observed for any patient in either opioid naive or exposed groups (Table 2).

**Table 2.**
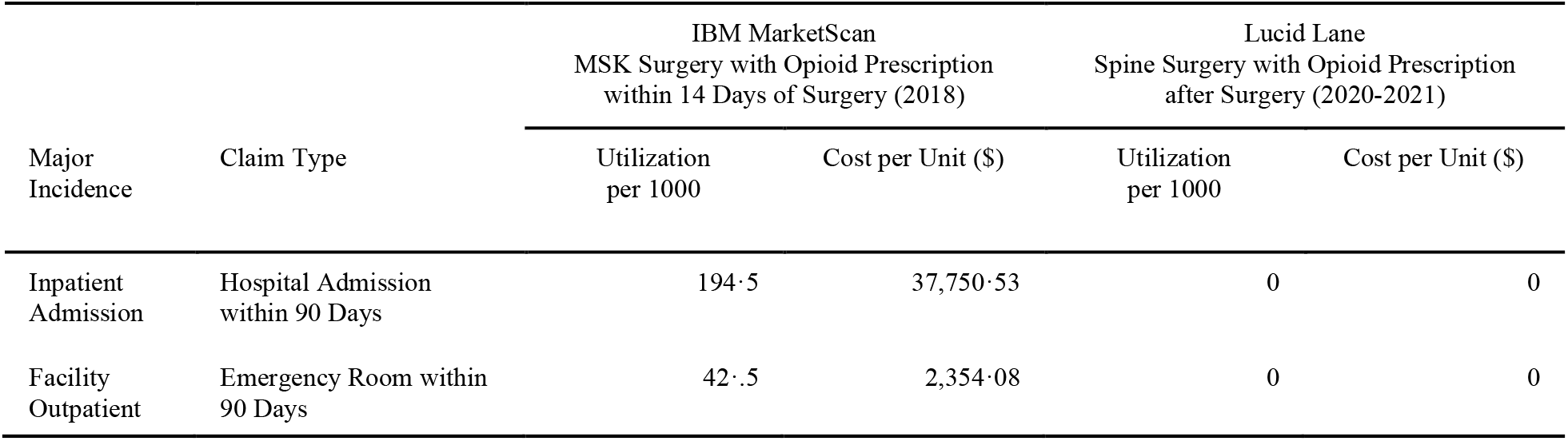
Comparison of healthcare utilization within 90-days postoperative period, IBM^R^Marketscan^R^ Data and Lucid Lane Data.

## Discussion

In this feasibility trial of 150 participants undergoing elective spine surgery who were enrolled into a virtual TPS with goals of opioid tapering and postoperative pain management facilitated through CBT delivered through a telehealth platform, we found that: i) engagement was high, ii) tapering to zero MMEs was achievable by all naive patients and most opioid exposed participants, iii) pain and other behavioral health scores improved significantly, and iv) no high-cost healthcare utilizations occurred, such as ER visits or rehospitalizations. Baseline socio-demographic, economic or clinical variables were not associated with any of the primary study outcomes. These results indicate that TPS facilitated by CBT-focused opioid tapering can be implemented virtually through telehealth with measurable benefits to postoperative patients.

Study engagement, defined as attendance in one or more post-surgical CBT-TPS sessions, was over 60% which is considerably higher than 20-40% reported engagement for telehealth studies among spine surgery patients.^33,34^ Of patients within the TPS at Toronto General Hospital, 41·3% engaged with the Manage My Pain mobile health platform.^35^ The higher levels of engagement seen in our study may be due to the structured nature of the program, which included clear preoperative expectation about the patient journey and explicit communication of opioid tapering as the goal defined by the CBT-focused protocol.

Approximately 13% of orthopedic spine surgery patients develop new persistent opioid use,^12^ making it a common post-surgical complication. Existing studies indicate that compared to preoperative use, 33% of opioid exposed patients increase, 33% decrease and 33% remain at the same MME postoperatively over a 12-month period.^36^ In our study, no participant increased their postoperative MME dosage. All engaged opioid naive participants (n = 67) tapered 100% off their postoperative opioids within 90 days (0 MME). These results compare favorably to an in-person TPS study in which, six months after surgery, only 46% (n = 51) of the opioid naive cohort tapered completely, and 8% (n = 9) actually increased their opioid usage.^21^ Seventy-four percent of opioid exposed participants reduced postoperative opioid MME by at least 50%. Interestingly, over half (53%) of exposed participants reduced their postoperative MME by 100%, thereby completely tapering off the opioid. These results also standout in comparison to an in-person TPS study in which, six months after surgery, 25% (n = 35) of opioid exposed patients had completely tapered off their postoperative opioids, and 19% (n = 27) increased their opioid usage.^21^

Opioids are often prescribed for postoperative analgesia. While the tapering results presented here are notable, it is unclear how they alone impact patients’ quality of life. We observed clinically and statistically significant improvements across all behavioral health symptoms, including depression, anxiety, QoL, and pain amongst both naive and exposed arms. These results are particularly important when viewed within the context of in-person TPS that did not utilize a CBT-focused protocol. While our study does not have matching inclusion criteria to the in-person TPS study at Toronto General Hospital, some comparison is warranted. Participants in our CBT-guided vTPS study experienced twice the improvement in pain symptoms, 5 times the reduction in depression, and 10 times the reduction in anxiety as was reported for patients in the landmark TPS study.^37^

The estimated economic costs of the US opioid epidemic topped $1 trillion in 2017.^38^ The largest drivers of direct healthcare costs associated with post-surgical opioid use are ER visits and re-hospitalizations, with these events occurring at a rate of over 400 per thousand patients (see Table 3).^17^ The fact that zero events occurred for any of the 96 patients completed this study is a promising indication that successful tapering while improving behavioral health could also potentially result in lower healthcare cost and utilization.^36^

While the results presented here are encouraging, the study has several limitations. The percentage of opioid exposed participants that completed the study was low. Patients with a history of opioid use for chronic pain are at risk of other surgical complications,^39^ and we saw several of these participants needing to leave the study because they required additional spine surgeries. While the outcomes observed here for exposed participants were statistically significant, a larger opioid exposed study size is needed in the future, including direct measures of the cost of implementing a program and subsequent economic benefit. The study was designed to test the feasibility of vTPS with greater incorporation of CBT-focused opioid tapering, so randomization was not used. Thus, we were unable to isolate the effect of improved pain and behavioral health over time in a non-therapy group. Further randomized studies are needed to evaluate vTPS over existing standards of care, although these promising findings may lead to ethical considerations if participants are required to be randomized to no therapy.

In summary, this study explored the feasibility of implementing a virtual TPS in a dual arm prospective trial of 150 opioid naive and opioid exposed participants from diverse backgrounds undergoing spinal surgery. Engagement was high, most participants tapered completely off opioids within 3 months regardless of study arm. Pain and behavioral health scores improved clinically, as well as statistically, and no high-cost opioid-related healthcare utilization events occurred. While traditional TPS programs are known to be effective for behavioral health, they may have limitations to greatly reduce postoperative opioid use and may be costly to implement. The evidence from this study indicates that a virtual TPS can offer significant health benefits while preventing new and persistent opioid use, and would likely be feasible for most hospitals to adopt.

## Methods

The Committee for the Protection of Human Subjects at the University of Texas Health Science Center Houston (IRB NUMBER: HSC-MS-20-0549) approved the study. All participants gave verbal and written consent to participate. The study was registered at clinicaltrials.gov, identifier NCT04787692. De-identified data will be provided upon request.

### Outcome measures

The primary outcomes were engagement with the CBT program after surgery, opioid tapering and improvements in behavioral health. We defined engagement as the participants attending at least one session with their assigned therapist after surgery.

We assessed opioid tapering by percent reduction from postoperatively prescribed opioids, which were standardized to morphine milligram equivalents (MME). For opioid naive participants, opioid tapering was assessed within 90 days after surgery and for opioid exposed participants, opioid taper was assessed within 180 days after surgery.

Participants reported pain, quality of life, functioning, general health symptoms, and anxiety and depression symptoms. We utilized validated, reliable survey instruments to assess these measures of behavioral health (See Supplementary Figure 1).

### Study Procedures

We recruited adult patients (aged 18 years or older) scheduled for elective orthopedic or neurosurgical spine surgery from the Anesthesia Preoperative Clinic at Memorial Hermann Hospital in the Texas Medical Center, the teaching facility associated with McGovern Medical School at UTHealth-Houston. Eligibility inclusion and exclusion criteria are shown in Supplementary Figure 2. After consent, the pre-surgery session was performed with collection of participants self-reported sociodemographic information and behavioral health surveys. Information regarding opioid exposure was cross referenced from the medical record.

At the time of study initiation, state mandated, COVID-19 hospital restrictions were implemented which led to interruption of scheduled elective surgery. In addition, for several periods, the pre-operative clinic functioned virtually with no in-patient visits. During these times, surgical co-investigators mentioned the study to subjects in preoperative visits and presented study details to subjects after arrival for their scheduled procedure by members of the research team. If the patient consented, enrollment and study procedures took place in a private room in the day surgery unit. Despite these unforeseen restrictions, study procedures continued as elective surgery was scheduled, and screening and enrollment resumed in-person once the pre-operative assessment clinic was opened.

Participants were scheduled for their initial therapy session, which was within seven days of discharge from the hospital, at the pre-operative screening. Therapy sessions occurred via HIPAA-compliant audio/video-calling platform, through a single-use link. Licensed therapists at Lucid Lane provided therapy services. Lucid Lane (Palo Alto, CA) is a telehealth company that focuses on administering CBT-facilitated tapering virtually (i.e., through HIPAA-compliant audio/video platforms) to perioperative patients, as well as other pain or general mental health patients. For the study, opioid naive participants received one one-hour session weekly for up to four weeks, to be used consecutively, after which they received a 90-day postoperative follow-up. At the first and last therapy sessions and at the follow-up, therapists asked participants to complete the behavioral health surveys and opioid use details (i.e., dosage and frequency of use). Therapists also tracked self-reported high-cost healthcare utilization events such as ER visits and re-hospitalizations. Opioid exposed participants received one one-hour session weekly for up to 26 weeks (180 days), to be used consecutively. If the patient dropped off weekly sessions prior to 26 weeks, the therapist conducted follow-up at 30-, 90- or 180-days post-operatively, depending on when the participants stopped attending therapy sessions. Participants were automatically removed from therapy sessions if they missed or could not be contacted for three consecutive appointments. There was no charge to participants for the therapy services.

A series of power analyses to compute achievable power were run using the Monte Carlo procedure in MPlus.^28,29^ We included variances based on preliminary data from existing Lucid Lane patients (rep corr = 0·3 to 0·5) and a nonsphericity correction ε = ·5, such that 1/[repetitions-1], while holding all other parameters constant. Due to high attrition rates (between 25 and 50%) from psychological interventions targeting treatment of opioid use, ^30^ we sought to consent n = 150 subjects, with anticipated subject fall-out of the postoperative therapy sessions, to detect medium effects (f = 0·25, α = 0·05). We conducted post-hoc sensitivity power analyses to identify power and detectable effect size based on the attrition samples. Based on a sample size n = 67 (opioid naïve), analyses indicated 80% power to detect at least small-to-medium effects (d = ·30, α = 0·05), and for n = 19 (opioid exposed), 80% power to detect at least medium-to-large effects (d = ·59, α = 0·05).

Lucid Lane therapists collected therapy data, with surgical and anesthesiology investigators blinded from these findings. We used IBM SPSS v 27 (Armonk, NY) to analyze: i) descriptive statistics, used for participants sociodemographic, pre-surgery and post-surgery behavioral health characteristics, ii) odds-ratio analyses, used to determine probability of engaging based on pre-surgery characteristics, iii) analysis of variance (ANOVA), used to determine differences in preoperative behavioral health scores based on opioid group (naïve vs exposed), iv) odds-ratio analyses, used to determine probability of meeting tapering goal based on pre-surgery characteristics and behavioral health, and v) Wilcoxon signed rank tests, used to determine changes in behavioral health from the first session to the final session. We report median values with interquartile ranges, which is best practice for clinical trial data that are skewed and cannot be normalized.^31,32^

We preserved participant confidentiality by storing paper informed consent forms and pre-surgical survey instruments in a locked office in the medical school building of the study PI. Furthermore, all electronic data were maintained on a HIPAA-compliant BOX folder.

## Author Contributions

MH: Lucid Lane PI, biostatistician, lead author, study design, administration of the study protocol, data maintenance and management, data analysis, initial complete manuscript draft, manuscript editing/reviewing.

BN: co-author, literature review, writing contributions to major sections of manuscript, manuscript feedback, editing/reviewing.

AZ: co-author, study design, manuscript feedback.

JS: co-author, access to resources/materials, manuscript feedback.

JC: co-author, access to resources/materials, manuscript feedback.

AF: co-author, access to resources/materials, manuscript feedback.

EP: UT Houston PI, study design, administration of the study protocol, access to resources/materials, manuscript feedback.

## Data Availability Statement

All data will be made available upon publication. It will be available in deidentified format.

